# Sedentary Behavior and Eating Habits Associated with Fast Food Consumption Among Bangladeshi University Students: The Moderating Role of Residential Status

**DOI:** 10.64898/2026.04.24.26351719

**Authors:** Abu Faisal, Mohammad Omar Nurshad, Taufiqul Alam, Anwoy Barua

## Abstract

**Objective:** To examine associations between sedentary behaviors, eating habits, and weekly fast food and sugar-sweetened beverage (SSB) consumption among university students in Bangladesh, and to assess whether residential status moderates these associations.

**Methods:** A cross-sectional study was conducted among 433 undergraduate and postgraduate students (aged 18–25 years) at the University of Chittagong, Bangladesh, using structured paper-based questionnaires. Bivariate associations were assessed using Pearson’s chi-square and Fisher’s exact tests. Two separate proportional odds models were fitted for fast food and SSB consumption, incorporating behavioral predictors and residential status interaction terms. The proportional odds assumption was verified using the Brant test.

**Results:** The overall prevalence of fast food consumption at least once per week was 91.88%. Frequent snacking (>3 days/week) between breakfast and lunch (OR = 2.17, p = 0.007), between lunch and dinner (OR = 1.77, p = 0.009), and after dinner (OR = 2.17, p = 0.043) was significantly associated with higher odds of frequent fast food consumption. Eating while watching TV or videos very often (OR = 2.68, p = 0.005) and consuming home-cooked meals (OR = 0.32, p = 0.005) were also significant predictors. Commuting more than 30 minutes daily was associated with higher odds of fast food intake (1–2 hours: OR = 2.00, p = 0.010). Among students in rented accommodation, each unit increase in emotional eating score was associated with 28% higher odds of more frequent fast food intake compared to dormitory residents (OR = 1.28, p = 0.034). For SSB consumption, snacking after dinner (OR = 2.55, p = 0.012), eating while watching TV or videos very often (OR = 2.75, p = 0.007), home-cooked meals (OR = 0.42, p = 0.035), and video gaming five or more hours daily (OR = 0.39, p = 0.002) were significant independent predictors.

**Conclusion:** Specific snacking behaviors, screen-associated eating, home-based meal access, and commuting time are independently associated with fast food and SSB consumption frequency among Bangladeshi university students. The moderating role of residential status on emotional eating suggests that dietary interventions should be tailored to students’ living arrangements.

## Introduction

Fast food consumption is rising globally, including in developing nations such as Bangladesh, where the fast food market generated approximately USD 1.75 billion in revenue in 2024 and is projected to reach around USD 3 billion by 2032, at a compound annual growth rate of approximately 7.1% [1]. Fast food is typically offered in restaurants and snack bars as a quick and inexpensive alternative to home-cooked meals [2]. However, it is often low in dietary fiber, micronutrients, and other essential nutrients but high in refined carbohydrates, sodium, saturated fats, trans fats, and cholesterol, making it a nutritionally poor dietary choice [3,4]. In Bangladesh, these health risks are further compounded by widespread food adulteration and poor hygiene practices, including the use of toxic artificial colors and harmful chemicals in food preparation [5].

Frequent fast food consumption has been strongly linked to adverse health outcomes, including obesity, type 2 diabetes, cardiovascular disease, and cognitive decline [6,7,8]. It is also associated with sleep disturbances, poor academic performance, and increased risk of mental health problems [9,10,11]. For example, children and adolescents in the highest quartile of fast food consumption had nearly three times greater odds of developing metabolic syndrome compared to the lowest quartile [12], and young women who consumed fast food more than twice per week demonstrated greater insulin resistance over a 15-year follow-up period [13].

Lifestyle factors have been consistently identified as correlates of fast food intake. Sedentary behavior, particularly screen-based activities, increases exposure to food advertising and is positively associated with fast food and soft drink consumption [14,15]. Skipping breakfast has also been found associated with fast food intake [16], that could lead to greater reliance on unhealthy food later in the day [17]. Eating late at night or while watching television has also been linked to fast food intake [18]. Time constraints, including prolonged commuting, have been shown to shift individuals toward convenient food options [19]. Taste preference and stress-driven eating further contribute to frequent fast food reliance among students [20,21].

University students represent a particularly vulnerable population. They experience increased autonomy, high academic demands, and shifting dietary environments. Objectively measured sedentary behavior in this group averages approximately 7.29 hours per day [22], with some estimates even higher [23]. In the Bangladeshi context, residential status may further shape dietary behaviors: among 53 public universities, 262 residential halls accommodate over 692,000 students [24]. Students living away from family may have limited access to nutritious home-cooked meals and may depend more heavily on convenient food options. Prior evidence from Bangladesh and Belgium suggests that residential students face greater dietary risk than non-residential peers [20,25].

Although growing evidence addresses psychological dimensions of fast food intake [11,26,27], few studies have jointly examined sedentary behavior and eating habits in relation to fast food consumption among South Asian university students. No prior study, to our knowledge, has assessed whether residential status moderates these behavioral associations. Therefore, this study aimed to examine associations between sedentary behavior, eating habits, and fast food consumption among university students in Bangladesh, and to assess the moderating role of residential status.

## Materials and Methods

### Study design and participants

This cross-sectional study enrolled currently registered undergraduate and postgraduate students at the University of Chittagong, Bangladesh. Eligible participants were aged 18 to 25 years and were actively attending academic activities during the data collection period. Students who did not provide written informed consent were excluded. A total of 433 students participated.

### Sample size

The required sample size was estimated using Cochran’s formula for proportions: n = Z² × p(1−p) / d², where Z = 1.96 (95% confidence level), p = 0.80 (higher than estimated fast food prevalence informed by prior studies [28,29]), and d = 0.05 (margin of error). This yielded a minimum of 246, adjusted to 273 after applying a 10% non-response correction. The achieved sample size of 433 exceeded this minimum.

### Sampling and data collection

Participants were recruited using a stratified convenience approach, with efforts made to include students from different academic departments, residential halls, and classroom settings to ensure representation across residential categories. Data was collected in August 2025 using a structured, paper-based questionnaire. A pilot study was conducted among 50 students prior to main data collection to assess the clarity, reliability, and feasibility of the instrument. Minor modifications were made to wording and structure based on pilot findings. Written informed consent was obtained from all participants before participation. Participation was entirely voluntary, and confidentiality of all responses was strictly maintained.

### Outcome variables

The primary outcome was weekly fast food consumption frequency, measured on a 4-point ordinal scale (rarely/never; 1–3 days/week; 4–6 days/week; daily). An ordinal rather than dichotomous scale was chosen to permit a more nuanced assessment of consumption patterns unlike prior studies [28,16]. Weekly sugar-sweetened beverage (SSB) intake was assessed using the same 4-point scale and analyzed in a separate ordinal model.

### Predictor and covariate variables

#### Sociodemographic factors

Gender (male, female), residential status (living with family; rented accommodation without family; university dormitory/hall), academic year (1st year through Master’s), personal monthly income from tuition or part-time employment (no income; <5,000 BDT; 5,000–10,000 BDT; >10,000 BDT), and receipt of family financial support (yes, no) were assessed. While previous studies often categorized financial status broadly (e.g., lower, middle & upper class [29]), this study used students’ own monthly income from tuition or part-time work for a more precise assessment.

#### Eating habits

Weekly frequency of breakfast, lunch, and dinner consumption was recorded and re-coded into binary categories (>3 days/week vs. ≤3 days/week). Snacking frequency was assessed across three intervals: between breakfast and lunch, between lunch and dinner, and after dinner, each also re-coded into the same binary categories. Dinner timing was categorized as before 9 PM, 9–10 PM, or after 10 PM. Primary meal location was categorized as home-cooked (self or family), university canteen or hall, or restaurants/food outlets. Frequency of eating while watching TV or videos was assessed on a 4-point scale (never, occasionally, often, very often).

#### Sedentary behaviors and physical activity

Sedentary time was assessed across multiple domains: attending academic classes, studying or completing assignments, reading non-academic books, using social media, playing video games, watching TV or streaming content, and daily commuting. Each domain was categorized into time intervals (<1 hour, 1–3 hours, 3–5 hours, ≥5 hours per day), with categories merged where necessary to ensure cell adequacy. Physical activity was assessed as frequency of structured exercise and sports participation, each categorized as rarely/never or at least once per week.

#### Sleep

Sleep duration has been found inversely related to fast food intake [16,30]. In our study, sleep duration was categorized as <7 hours, 7–9 hours, or >9 hours per night as current consensus recommendations suggest that adults should obtain at least 7 hours of sleep per night to maintain optimal health [31]. Sleep onset time was also assessed & categorized as before midnight or after midnight.

#### Composite scores

An emotional eating composite score was derived from three items assessing eating in response to stress, boredom, and anger, each rated on a 4-point frequency scale (rarely/never to daily). This approach is inspired by established multi-item emotional eating assessments [32], with acceptable internal consistency (Cronbach’s α = 0.637). A screen time composite score was computed as the aggregate of sedentary time spent on social media and watching TV or streaming (Cronbach’s α = 0.576), following an approach validated by Fletcher et al. [33]. Video gaming time was retained as a separate categorical predictor because its inclusion in the composite adversely reduced the scale’s reliability.

### Statistical analysis

Descriptive statistics summarized the distribution of participant characteristics across ordinal consumption categories. Bivariate associations between each predictor and each outcome were assessed using Pearson’s chi-square test; Fisher’s exact test was applied when expected cell counts fell below 5.

Two separate proportional odds (ordinal logistic regression) models were constructed, one for fast food consumption and one for SSB consumption. Predictors were included in the multivariable model if they demonstrated a statistically significant bivariate association (p < 0.05) with the relevant outcome. Composite scores were entered as continuous covariates.

Interaction terms between residential status and each composite score were included to evaluate potential moderation; variables requiring Fisher’s exact test in bivariate analysis were excluded from interaction testing to avoid sparse data issues and quasi-complete separation. The proportional odds assumption was verified using the Brant test (Test of Parallel Lines). Statistical significance was set at α = 0.05. All analyses were performed using IBM SPSS Statistics version 25 and RStudio version 4.1.3.

### Ethical approval

Ethical approval was obtained from the Institutional Review Board of the Bangladesh Institute of Innovative Health Research (reference number: BIIHR-2025-021). All procedures were conducted in accordance with the Declaration of Helsinki.

## Results

### Participant characteristics and bivariate findings

A total of 433 students participated (males: 66.4%; females: 33.6%). The overall prevalence of fast food consumption at least once per week was 91.9%. Frequent consumption (>3 days/week) was reported by 74.60% for fast food and 90.26% for SSBs (Fig 1). No statistically significant association was observed between any sociodemographic variable—including gender (p = 0.917) and residential status (p = 0.055)—and fast food or SSB intake in bivariate analysis (Table 1).

**Fig 1.**
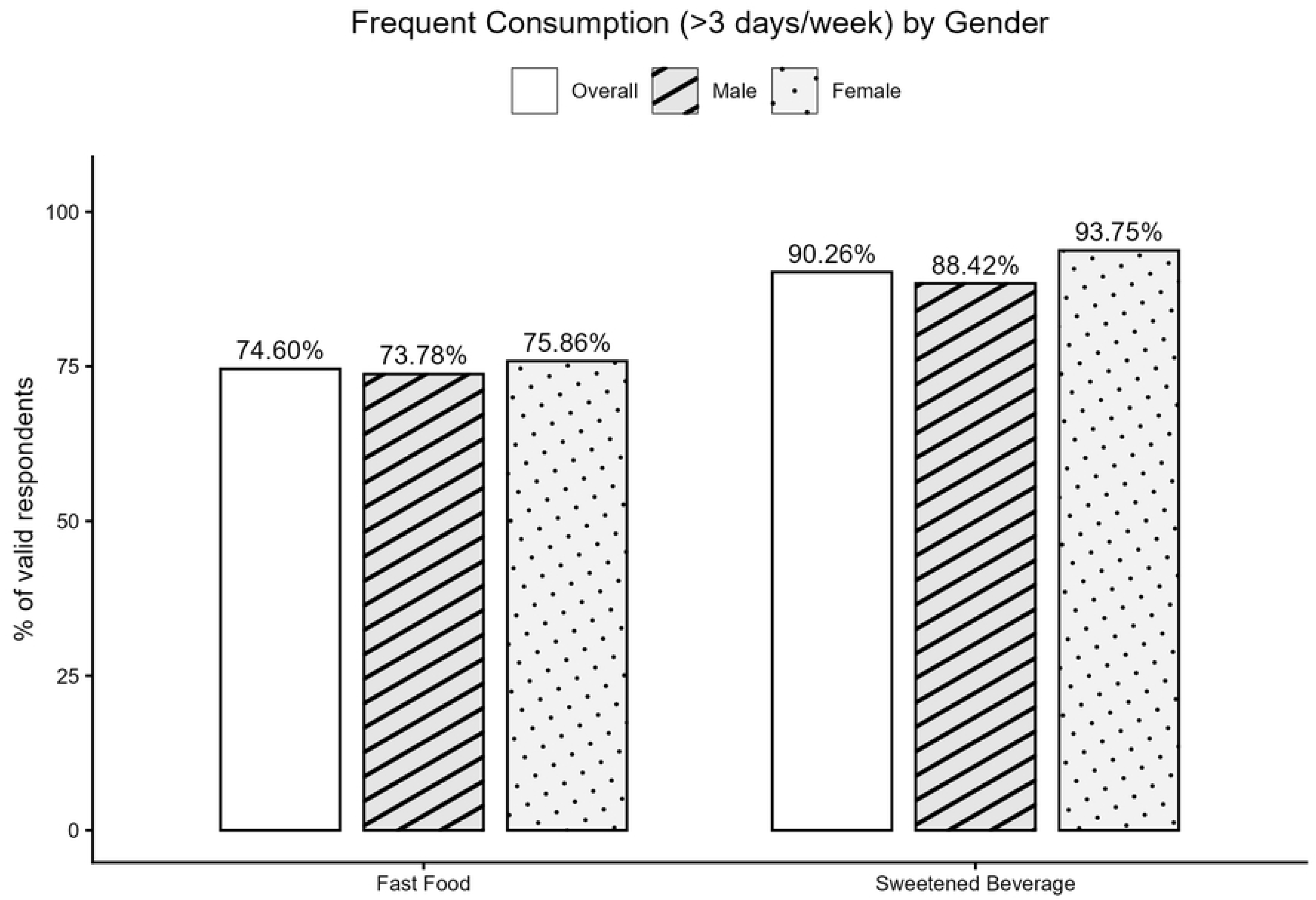
Prevalence of frequent consumption of fast food and sugar-sweetened beverages across gender.

Despite the absence of statistical significance, bivariate patterns by residential status were notable. Students in rented accommodation had the highest proportion of frequent fast food consumers (77.93%) (Fig 2), while those living with family had the highest SSB consumption frequency (93.55%). Dormitory residents had the lowest proportions for both outcomes (fast food: 68.99%; SSB: 85.35%).

**Fig 2.**
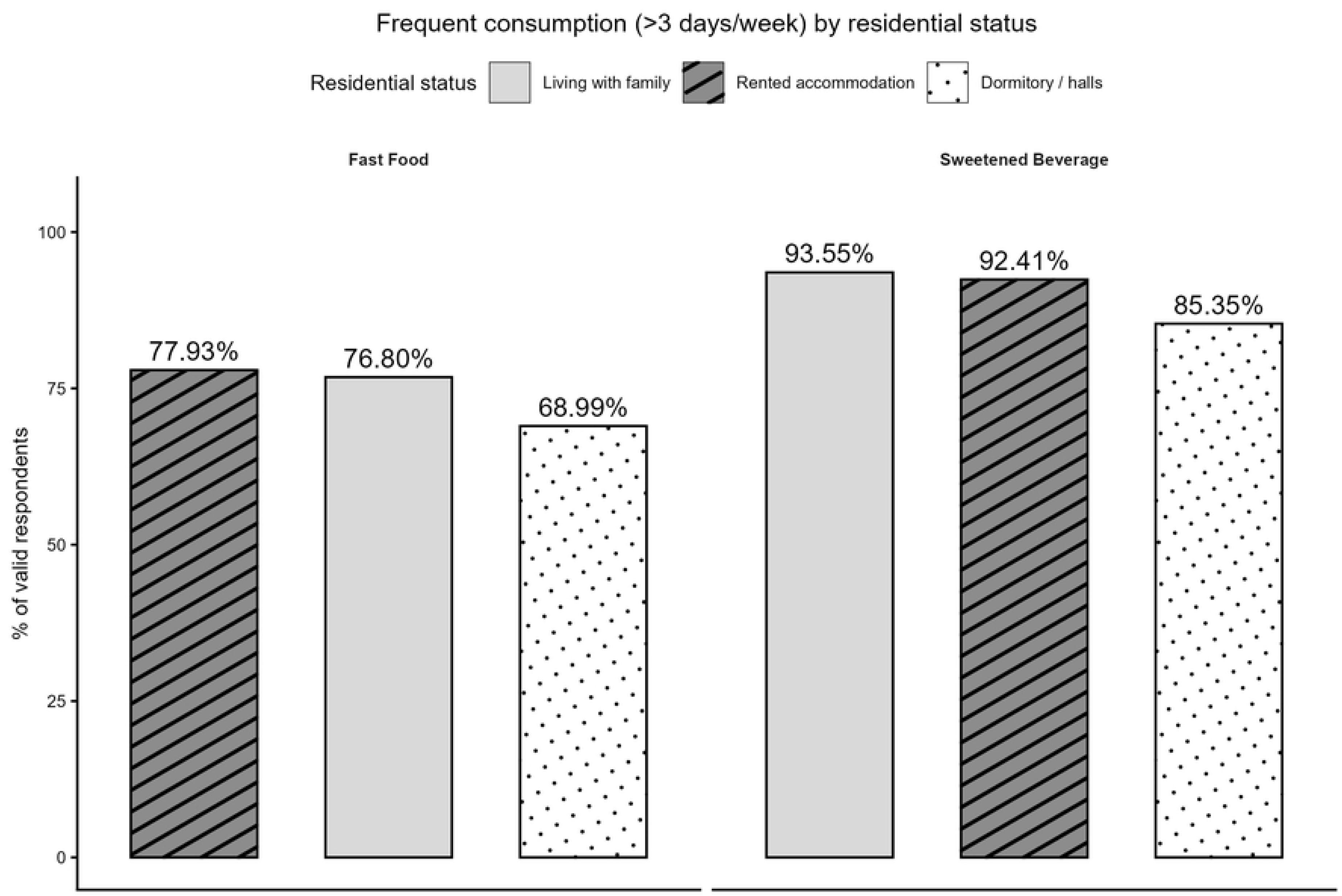
Prevalence of frequent consumption of fast food and sugar-sweetened beverages across residential status.

Six eating behaviors and three sedentary behaviors were significantly associated with both fast food and SSB intake (Tables 2 and 3). Breakfast frequency (p < 0.01 for both), snacking between lunch and dinner (p < 0.05 for both), snacking after dinner (p < 0.01 for both), eating while bored (fast food: p = 0.005; SSB: p = 0.010), eating while watching TV or videos (fast food: p = 0.008; SSB: p = 0.019), and primary meal location (fast food: p = 0.016; SSB: p = 0.001) were each significantly associated with both outcomes. Sedentary time spent studying (fast food: p = 0.018; SSB: p = 0.012), using social media (p < 0.01 for both), and playing video games (p < 0.01 for both) were also significantly associated with both outcomes.

Associations exclusive to fast food included dinner frequency (p = 0.037), snacking between breakfast and lunch (p = 0.001), dinner timing (p = 0.018), commuting time (p = 0.042), and sleep onset time (p = 0.005). An association with eating when stressed was exclusive to SSB consumption (p = 0.016).

### Proportional odds model for fast food consumption

The model demonstrated acceptable fit (model fitting information: p < 0.001; Pearson goodness-of-fit: p = 0.116; McFadden’s pseudo R² = 0.113) and the proportional odds assumption was not violated (Test of Parallel Lines: p = 0.238).

Snacking more than three days per week was significantly associated with higher odds of frequent fast food intake at all three intervals: between breakfast and lunch (OR = 2.17, 95% CI: 1.24–3.79, p = 0.007), between lunch and dinner (OR = 1.77, 95% CI: 1.16–2.71, p = 0.009), and after dinner (OR = 2.17, 95% CI: 1.03–4.58, p = 0.043). Eating while watching TV or videos was a significant predictor when reported very often (OR = 2.68, 95% CI: 1.34–5.33, p = 0.005) and often (OR = 2.33, 95% CI: 1.19–4.55, p = 0.013). Consuming home-cooked meals was associated with substantially lower odds of frequent fast food intake compared to eating at restaurants or food outlets (OR = 0.32, 95% CI: 0.14–0.72, p = 0.005).

Commuting time greater than 30 minutes daily was significantly associated with higher odds of fast food intake. Compared with students commuting fewer than 30 minutes, those commuting 1–2 hours had the highest odds (OR = 2.00, 95% CI: 1.18–3.39, p = 0.010), followed by those commuting more than 2 hours (OR = 1.94, 95% CI: 1.04–3.61, p = 0.037) and 30 minutes to 1 hour (OR = 1.81, 95% CI: 1.05–3.13, p = 0.033). A significant moderation effect was found for students living in rented accommodation without family: each unit increase in emotional eating score was associated with 28% higher odds of more frequent fast food consumption compared to dormitory residents (OR = 1.28, 95% CI: 1.02–1.60, p = 0.034). Residential status, breakfast and dinner frequency, sedentary study time, video gaming duration, sleep onset time, and composite screen time were not significant independent predictors after adjustment.

**Table 4:** Proportional odds model for weekly fast food consumption frequency (n=433)

| Variable | OR | p | 95% CI<br>Lower | 95% CI<br>Upper |
| --- | --- | --- | --- | --- |
| Residential status: living with family | 0.38 | 0.256 | 0.07 | 2.03 |
| Residential status: rented accommodation (without family) | 0.29 | 0.128 | 0.06 | 1.42 |
| Residential status: university dormitory/halls [Ref] | — | — | — | — |
| Breakfast frequency: >3 days/week | 0.85 | 0.485 | 0.53 | 1.35 |
| Breakfast frequency: ≤3 days/week [Ref] | — | — | — | — |
| Dinner frequency: >3 days/week | 1.37 | 0.469 | 0.59 | 3.20 |
| Dinner frequency: ≤3 days/week [Ref] | — | — | — | — |
| Snack BL: >3 days/week | 2.17 | 0.007 | 1.24 | 3.79 |
| Snack BL: ≤3 days/week [Ref] | — | — | — | — |
| Snack LD: >3 days/week | 1.77 | 0.009 | 1.16 | 2.71 |
| Snack LD: $\leq 3$ days/week [Ref] | — | — | — | — |
| Snack after dinner: $>3$ days/week | 2.17 | 0.04<br>3 | 1.03 | 4.58 |
| Snack after dinner: $\leq 3$ days/week [Ref] | — | — | — | — |
| Eat while watching TV/videos: Very often | 2.68 | 0.00<br>5 | 1.34 | 5.33 |
| Eat while watching TV/videos: Often | 2.33 | 0.01<br>3 | 1.19 | 4.55 |
| Eat while watching TV/videos: Occasionally | 1.73 | 0.10<br>5 | 0.89 | 3.37 |
| Eat while watching TV/videos: Never [Ref] | — | — | — | — |
| Dinner time: after 10 PM | 0.52 | 0.05<br>4 | 0.27 | 1.01 |
| Dinner time: 9–10 PM | 0.77 | 0.26<br>8 | 0.48 | 1.23 |
| Dinner time: before 9 PM [Ref] | — | — | — | — |
| Main meal location: home-cooked | 0.32 | 0.00<br>5 | 0.14 | 0.72 |
| Main meal location: university canteen/hall | 0.76 | 0.47<br>9 | 0.36 | 1.61 |
| Main meal location: restaurants/food outlets [Ref] | — | — | — | — |
| Studying $\geq 5$ hours/day | 1.15 | 0.60<br>4 | 0.67 | 1.99 |
| Studying 3–5 hours/day | 0.87 | 0.55<br>9 | 0.54 | 1.40 |
| Studying $<3$ hours/day [Ref] | — | — | — | — |
| Video gaming $\geq 5$ hours/day | 0.75 | 0.32<br>8 | 0.42 | 1.34 |
| Video gaming 3–5 hours/day | 1.74 | 0.12<br>4 | 0.86 | 3.51 |
| Video gaming $<3$ hours/day [Ref] | — | — | — | — |
| Commuting >2 hours/day | 1.94 | 0.03<br>7 | 1.04 | 3.61 |
| Commuting 1–2 hours/day | 2.00 | 0.01<br>0 | 1.18 | 3.39 |
| Commuting 30 min–1 hour/day | 1.81 | 0.03<br>3 | 1.05 | 3.13 |
| Commuting <30 min/day [Ref] | — | — | — | — |
| Sleep onset: after midnight | 1.54 | 0.08<br>8 | 0.94 | 2.52 |
| Sleep onset: before midnight [Ref] | — | — | — | — |
| Emotional eating composite score | 0.93 | 0.39<br>4 | 0.79 | 1.10 |
| Screen time composite score (social media + TV/streaming) | 1.04 | 0.68<br>9 | 0.85 | 1.27 |
| Living with family × emotional eating | 1.22 | 0.09<br>1 | 0.97 | 1.53 |
| Rented accommodation × emotional eating | 1.28 | 0.03<br>4 | 1.02 | 1.60 |
| Dormitory × emotional eating [Ref] | — | — | — | — |
| Living with family × screen time | 1.12 | 0.43<br>8 | 0.84 | 1.48 |
| Rented accommodation × screen time | 1.12 | 0.41<br>1 | 0.85 | 1.47 |
| Dormitory × screen time [Ref] | — | — | — | — |

*BL = between breakfast and lunch; LD = between lunch and dinner. Reference categories are indicated by [Ref]. Significant associations ( $p < 0.05$ ) are in bold. Model fitting: $p < 0.001$ ; Pearson goodness-of-fit: $p = 0.116$ ; McFadden's pseudo $R^2 = 0.113$ ; Test of Parallel Lines: $p = 0.238$ .*

### Proportional odds model for SSB consumption

The SSB model showed good overall fit (model fitting information: p < 0.001; Pearson goodness-of-fit: p = 0.906; McFadden’s pseudo R² = 0.077) and satisfied the proportional odds assumption (Test of Parallel Lines: p = 0.881).

Snacking after dinner more than three days per week (OR = 2.55, 95% CI: 1.22–5.31, p = 0.012) and eating while watching TV or videos very often (OR = 2.75, 95% CI: 1.32–5.73, p = 0.007) were significantly associated with higher odds of frequent SSB intake. Consuming home-cooked meals was associated with lower odds (OR = 0.42, 95% CI: 0.19–0.94, p = 0.035). Spending five or more hours per day playing video games was associated with significantly lower odds of frequent SSB intake (OR = 0.39, 95% CI: 0.22–0.71, p = 0.002). Each unit increase in emotional eating score was associated with higher odds of frequent SSB intake (OR = 1.23, 95% CI: 1.04–1.46, p = 0.017). No significant interaction effects between residential status and composite scores were observed for SSB consumption.

**Table 5.**
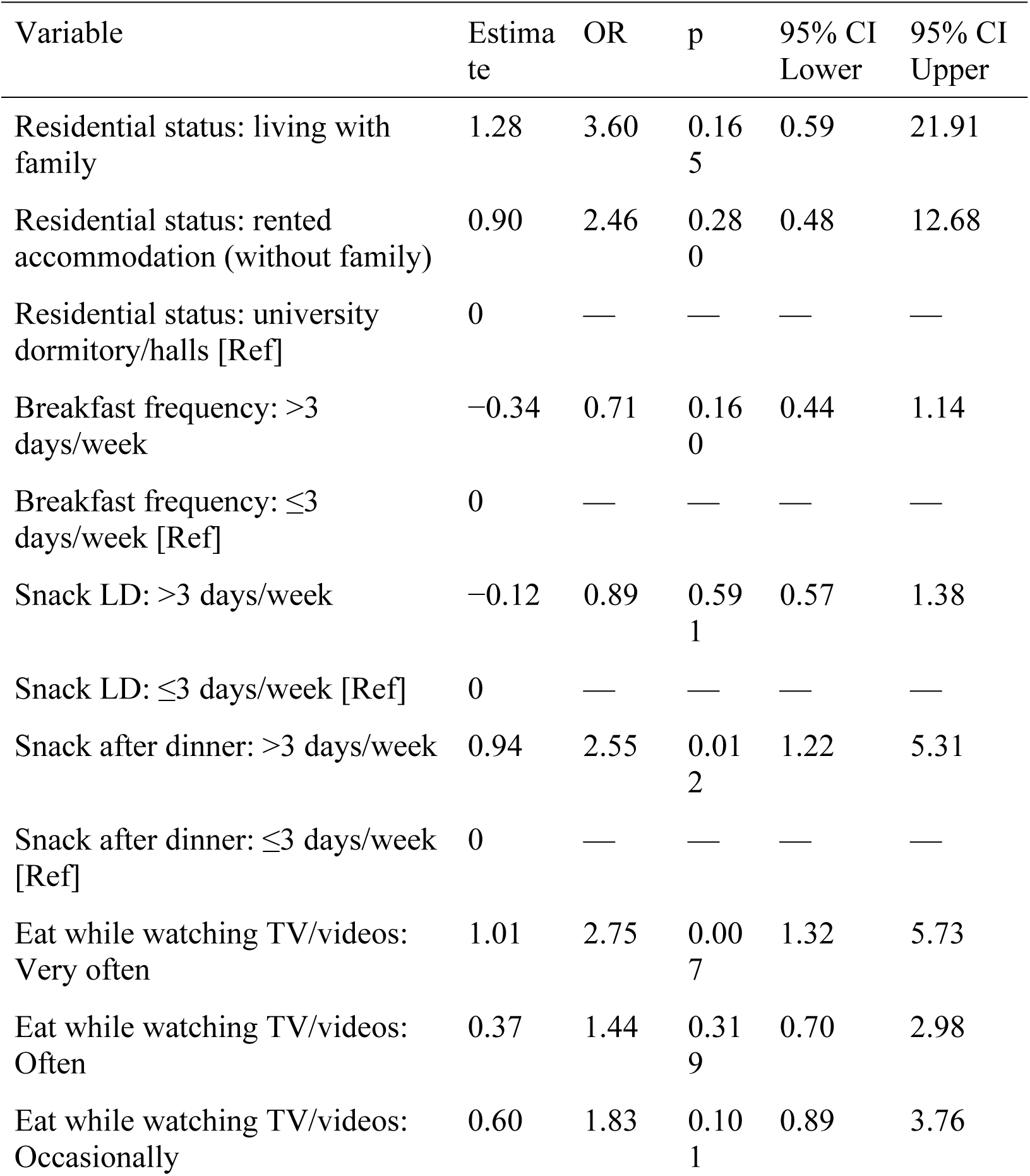
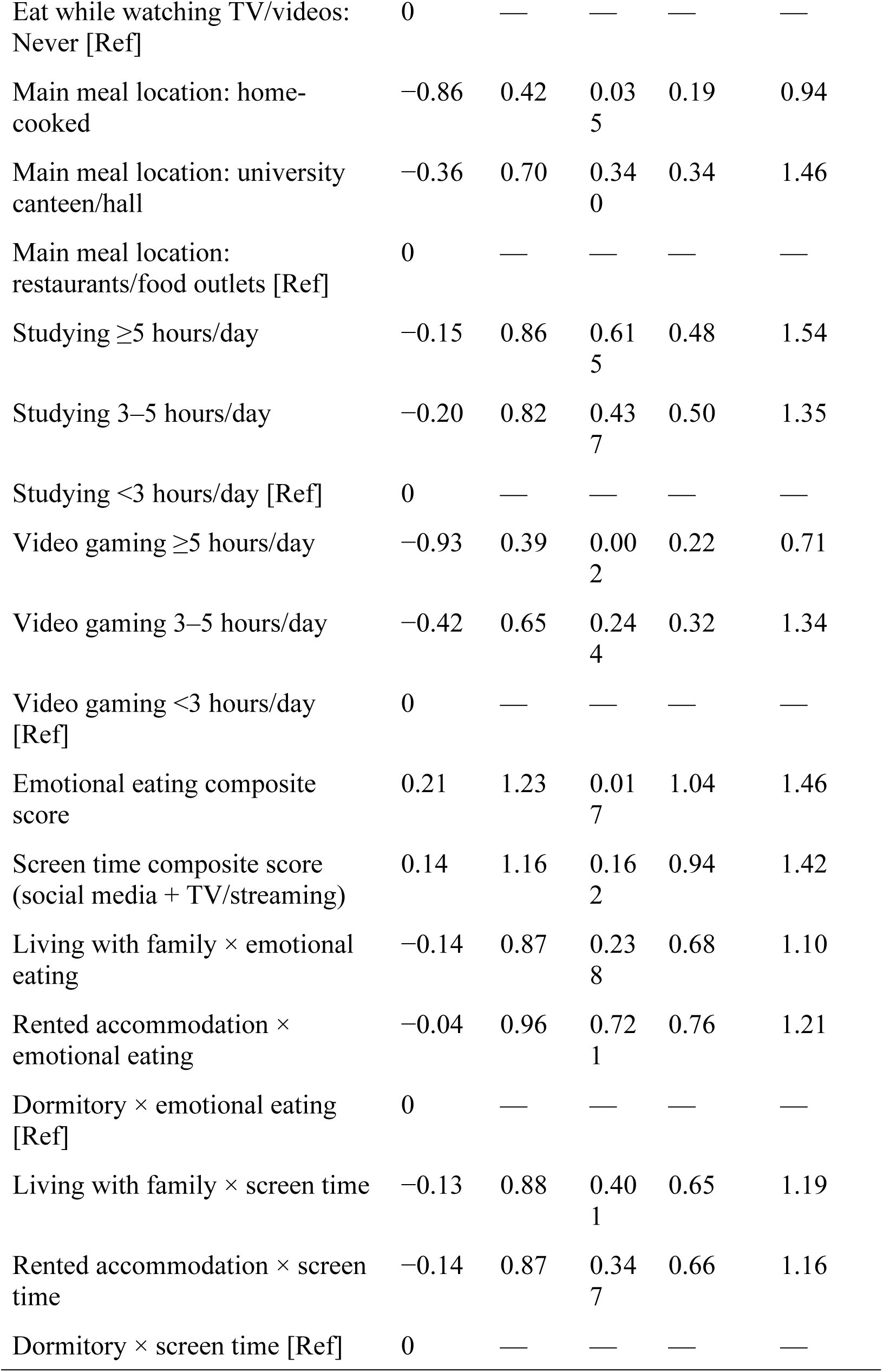
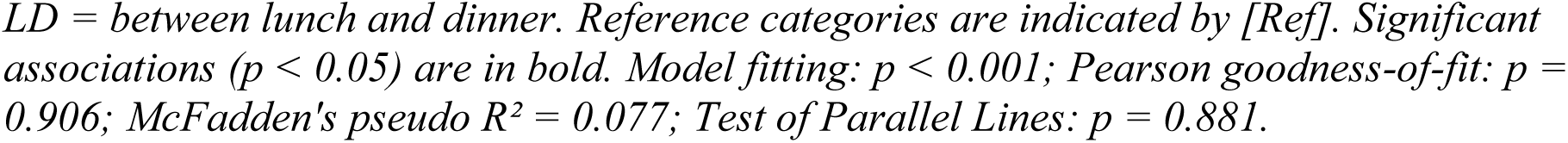
Proportional odds model for weekly sugar-sweetened beverage consumption frequency (n =431).

| Variable | Estimate | OR | p | 95% CI Lower | 95% CI Upper |
| --- | --- | --- | --- | --- | --- |
| Residential status: living with family | 1.28 | 3.60 | 0.165 | 0.59 | 21.91 |
| Residential status: rented accommodation (without family) | 0.90 | 2.46 | 0.280 | 0.48 | 12.68 |
| Residential status: university dormitory/halls [Ref] | 0 | — | — | — | — |
| Breakfast frequency: >3 days/week | −0.34 | 0.71 | 0.160 | 0.44 | 1.14 |
| Breakfast frequency: ≤3 days/week [Ref] | 0 | — | — | — | — |
| Snack LD: >3 days/week | −0.12 | 0.89 | 0.591 | 0.57 | 1.38 |
| Snack LD: ≤3 days/week [Ref] | 0 | — | — | — | — |
| Snack after dinner: >3 days/week | 0.94 | 2.55 | 0.012 | 1.22 | 5.31 |
| Snack after dinner: ≤3 days/week [Ref] | 0 | — | — | — | — |
| Eat while watching TV/videos: Very often | 1.01 | 2.75 | 0.007 | 1.32 | 5.73 |
| Eat while watching TV/videos: Often | 0.37 | 1.44 | 0.319 | 0.70 | 2.98 |
| Eat while watching TV/videos: Occasionally | 0.60 | 1.83 | 0.101 | 0.89 | 3.76 |
| Eat while watching TV/videos:<br>Never [Ref] | 0 | — | — | — | — |
| Main meal location: home-cooked | −0.86 | 0.42 | 0.03<br>5 | 0.19 | 0.94 |
| Main meal location: university canteen/hall | −0.36 | 0.70 | 0.34<br>0 | 0.34 | 1.46 |
| Main meal location:<br>restaurants/food outlets [Ref] | 0 | — | — | — | — |
| Studying $\geq 5$ hours/day | −0.15 | 0.86 | 0.61<br>5 | 0.48 | 1.54 |
| Studying 3–5 hours/day | −0.20 | 0.82 | 0.43<br>7 | 0.50 | 1.35 |
| Studying $< 3$ hours/day [Ref] | 0 | — | — | — | — |
| Video gaming $\geq 5$ hours/day | −0.93 | 0.39 | 0.00<br>2 | 0.22 | 0.71 |
| Video gaming 3–5 hours/day | −0.42 | 0.65 | 0.24<br>4 | 0.32 | 1.34 |
| Video gaming $< 3$ hours/day<br>[Ref] | 0 | — | — | — | — |
| Emotional eating composite score | 0.21 | 1.23 | 0.01<br>7 | 1.04 | 1.46 |
| Screen time composite score<br>(social media + TV/streaming) | 0.14 | 1.16 | 0.16<br>2 | 0.94 | 1.42 |
| Living with family × emotional eating | −0.14 | 0.87 | 0.23<br>8 | 0.68 | 1.10 |
| Rented accommodation ×<br>emotional eating | −0.04 | 0.96 | 0.72<br>1 | 0.76 | 1.21 |
| Dormitory × emotional eating<br>[Ref] | 0 | — | — | — | — |
| Living with family × screen time | −0.13 | 0.88 | 0.40<br>1 | 0.65 | 1.19 |
| Rented accommodation × screen time | −0.14 | 0.87 | 0.34<br>7 | 0.66 | 1.16 |
| Dormitory × screen time [Ref] | 0 | — | — | — | — |

*LD = between lunch and dinner. Reference categories are indicated by [Ref]. Significant* *associations ( $p < 0.05$ ) are in bold. Model fitting: $p < 0.001$ ; Pearson goodness-of-fit: $p =$* *0.906; McFadden's pseudo $R^2 = 0.077$ ; Test of Parallel Lines: $p = 0.881$ .*

## Discussion

This cross-sectional study examined behavioral and sedentary correlates of fast food and SSB consumption among university students at the University of Chittagong, with a specific focus on the moderating role of residential status. Frequent snacking, screen-associated eating, home-based meal access, and daily commuting time were independently associated with fast food intake, while residential status moderated the effect of emotional eating on fast food consumption. Several similar behavioral predictors also emerged for SSB intake.

### Prevalence and gender

The overall prevalence of fast food consumption at least once per week was 91.9%, substantially higher than the 82.9% reported by Islam et al. [29], though direct comparisons are limited by differences in measurement instruments and definitions. Applying a threshold of more than three days per week, the prevalence of frequent consumption was 74.6%, exceeding the 64% reported among Bangladeshi college students [28]. No significant gender difference was observed in the present study, which is inconsistent with prior studies identifying male gender as a predictor of higher fast food intake [16,18,21]. This discrepancy may reflect specific features of the university environment studied, shifting gender norms in dietary behaviors among urban Bangladeshi young adults, or sample composition.

### Snacking

Frequent snacking across all three daytime intervals was positively associated with higher odds of fast food consumption. This pattern is consistent with the notion that habitual snacking behavior may reflect a general tendency toward opportunistic food consumption, including fast food. Among previous studies, Fletcher et al. [33] did not directly link snacking to fast food intake but associated frequent discretionary snacking with higher screen time and sedentary behavior, suggesting possible indirect pathways. Snacking after dinner was also associated with higher SSB intake, indicating that late-evening eating occasions may involve both solid fast food and sweetened beverages.

### Eating while watching TV or videos

Eating while watching TV or videos was significantly associated with higher odds of both fast food (OR = 2.68) and SSB consumption (OR = 2.75) when done very often. This finding is consistent with evidence that university students with high adherence to the ‘fast food dominated’ dietary pattern, are more likely to eat while watching television [18]. Prior work reported that two or more hours of daily screen time was associated with higher fast food intake [33]. In the present study, composite screen time was not a significant independent predictor in either model; however, the specific behavior of eating while watching TV—assessed separately—emerged as a robust predictor for both outcomes.

### Home-cooked meals

Consuming home-cooked meals as the primary meal source was consistently associated with lower odds of frequent fast food and SSB consumption relative to eating at restaurants or food outlets. This finding is consistent with evidence that reliance on external food environments promotes fast food intake [20,25] and supports the importance of home food access as a protective dietary factor.

### Commuting time

Longer daily commuting time was significantly associated with higher odds of fast food intake across all categories above 30 minutes per day, consistent with prior evidence that time constraints shift food choices toward convenience options [19,25]. Students who spend considerable time commuting may have reduced opportunities to prepare or access home-cooked meals, increasing reliance on fast food vendors encountered along travel routes.

### Residential status and emotional eating

The significant interaction between rented accommodation and emotional eating (OR = 1.28, p = 0.034) indicates that emotional eating exerts a stronger positive association with fast food consumption among students living away from family in rented housing compared to dormitory residents. Students in rented accommodation may experience greater autonomy, social isolation, and fewer structural eating routines, which may amplify the role of emotional triggers in food decisions. This contrasts with dormitory residents, who may benefit from shared eating environments that buffer impulsive or emotional eating. This finding underscores the importance of considering students’ living contexts when designing dietary interventions. The positive effect of emotional eating on fast food intake is supported by a prior, large, longitudinal study as it reported that school going students with high emotional eating had higher odds of consuming fast food (OR = 2.40) [34].

### Physical exercise finding

Physical exercise and sports participation were not significantly associated (neither with fast food nor SSB intake) in bivariate analysis and thereby not included as predictors in the ordinal logistic regression models. In contrast, Tambalis et al. reported significant association and direct effect on fast food intake (among school going students aged not more than 18 years) [16]. Tareq et al. focused on obesity level rather than fast-food intake and found no significant association (in bivariate analysis) between BMI and frequency of participating in sports or physical exercise [35]. Notably, a very small proportion of participants in our study engaged in physical exercise or sports at least once per week. Future studies could adopt a more balanced approach like case control study to ensure better understanding of this relationship.

### Video gaming

Spending five or more hours per day playing video games was associated with significantly lower odds of frequent SSB consumption (OR = 0.39, p = 0.002) in the SSB model, while it was not a significant predictor for fast food intake. This unexpected finding may reflect the longer stay at home due to prolonged gaming sessions and thus leading to the dependence on home cooked meals as primary meal source. One finding in our study is that among the students who consumed home cooked meals as their main meal, 72.5% spent 5 hours or more daily playing video games.

### Sleep timing

Although sleep onset after midnight was significantly associated with fast food intake in bivariate analysis (p = 0.005), it did not independently predict fast food or SSB consumption in the multivariable models. This suggests that sleep timing may be confounded by other behavioral factors, though it may still be relevant to dietary patterns and merits examination in larger, longitudinal designs.

## Limitations

Several limitations of this study must be acknowledged. First, the cross-sectional design precludes causal inference; observed associations cannot establish directionality, and reverse causality cannot be excluded. Second, participants were recruited using a stratified convenience approach from a single public university, which limits the generalizability of findings to other institutions or populations. Third, all measurements were based on self-report, which is susceptible to recall bias, social desirability bias, and measurement error, particularly for dietary and physical activity variables. Fourth, certain subgroups were small (e.g., the category of students spending fewer than seven hours sleeping included approximately 18 individuals), reducing the statistical power and reliability of estimates within those cells. Fifth, the reliability of the composite scores was modest (emotional eating: Cronbach’s α = 0.637; screen time: Cronbach’s α = 0.576), and the screen time composite did not capture all potentially relevant sedentary domains. Sixth, interaction testing was restricted to composite scores to avoid model overfitting and quasi-complete separation, which may have prevented the detection of moderation effects involving other predictor categories.

## Conclusions

This study identified specific eating behaviors and commuting time as significant correlates of fast food and SSB consumption frequency among Bangladeshi university students. The finding that residential status moderates the association between emotional eating and fast food intake highlights the role of the living environment in shaping dietary behavior. Dietary health promotion programs targeting university students should incorporate context-specific strategies that account for students’ residential arrangements and daily behavioral patterns. Longitudinal and multi-site research is needed to establish directionality and improve generalizability.

## Data Availability

All data underlying the findings of this study are available in the paper and its Supporting Information files. The de-identified dataset is additionally available in CSV format on Figshare under a Creative Commons Attribution 4.0 International licence (CC BY 4.0) at https://doi.org/10.6084/m9.figshare.32025564.

## Acknowledgments

The authors thank all students who voluntarily participated in this study and specially thank Nazmul Hasan, Junayed Mohammed and Miskat Maruf for their support during data collection.

## Author Contributions

Md. Abu Faisal [M.A.F]: Conceptualization, Methodology, Formal analysis, Data curation, Writing – original draft, Preliminary Analysis, Writing – review & editing.

Mohammad Omar Nurshad [M.O.N]: Conceptualization, Investigation, Statistical Analysis, Data curation, Writing – original draft, Writing – review & editing.

Md. Taufiqul Alam [M.T.A]: Investigation, Data curation, Statistical Analysis, literature review, Writing – review & editing.

Anwoy Barua [A.B.]: Investigation, Data curation, Writing – review & editing.

## Competing Interests

The authors declare that they have no competing interests.

## Funding

This research received no specific grant from any funding agency in the public, commercial, or not-for-profit sectors. The authors received no financial support for the research, authorship, or publication of this article.

## Supporting Information

S1 Table. Full bivariate distribution tables for all study variables. (DOCX)

S2 File. Supplementary analyses, including threshold parameters, complete non-significant predictor estimates, video gaming and sports results with confidence interval width explanation. (DOCX)

S3 File. R analysis script for Fisher’s Exact test & plots reported in this study. (ZIP)

S4 File. Full SPSS statistical output for all analyses reported in the manuscript. Includes custom cross-tabulation tables, chi-square test results, proportional odds model parameter estimates, goodness-of-fit statistics, and test of parallel lines for both the fast food and sugar-sweetened beverage models. (DOCX)

Data availability statement

All relevant data are within the paper and its Supporting Information files at Figshare (doi:10.6084/m9.figshare.32025564).

The de-identified dataset is available in CSV format under a Creative Commons Attribution 4.0 International licence (CC BY 4.0). The complete questionnaire is separately provided under supporting information.

## Notes

### Competing Interest Statement

The authors have declared no competing interest.

### Funding Statement

The author(s) received no specific funding for this work.

### Author Declarations

This study was approved by the Institutional Review Board of the Bangladesh Institute of Innovative Health Research, Dhaka, Bangladesh (approval reference number: BIIHR-2025-021). All study procedures were conducted in accordance with the ethical principles outlined in the Declaration of Helsinki. Written informed consent was obtained from all participants prior to their inclusion in the study.

## References

1. Bangladesh Quick Service Restaurants Market Size, Share & Trends Analysis, 2032. In: P&S Intelligence [Internet]. [cited 20 Apr 2026]. Available: https://www.psmarketresearch.com/market-analysis/bangladesh-qsr-market-report

2. Fast food: MedlinePlus Medical Encyclopedia Image. [cited 20 Apr 2026]. Available: https://medlineplus.gov/ency/imagepages/19491.htm

3. Bahadoran Z, Mirmiran P, Azizi F. Fast Food Pattern and Cardiometabolic Disorders: A Review of Current Studies. Health Promot Perspect. 2016;5: 231–240. doi:10.15171/hpp.2015.028

4. Li L, Sun N, Zhang L, Xu G, Liu J, Hu J, et al. Fast food consumption among young adolescents aged 12–15 years in 54 low- and middle-income countries. Glob Health Action. 13: 1795438. doi:10.1080/16549716.2020.1795438

5. Food adulteration and inadequate hygiene practices endangering public health in Bangladesh | Discover Food | Springer Nature Link. [cited 20 Apr 2026]. Available: https://link.springer.com/article/10.1007/s44187-024-00191-8

6. Alsabieh M, Alqahtani M, Altamimi A, Albasha A, Alsulaiman A, Alkhamshi A, et al. Fast food consumption and its associations with heart rate, blood pressure, cognitive function and quality of life. Pilot study. Heliyon. 2019;5: e01566. doi:10.1016/j.heliyon.2019.e01566

7. Mazidi M, Speakman JR. Association of Fast-Food and Full-Service Restaurant Densities With Mortality From Cardiovascular Disease and Stroke, and the Prevalence of Diabetes Mellitus. J Am Heart Assoc. 2018;7: e007651. doi:10.1161/JAHA.117.007651

8. Payab M, Kelishadi R, Qorbani M, Motlagh ME, Ranjbar SH, Ardalan G, et al. Association of junk food consumption with high blood pressure and obesity in Iranian children and adolescents: the CASPIAN-IV Study. J Pediatr (Rio J). 2015;91: 196–205. doi:10.1016/j.jped.2014.07.006

9. Khan A, Dix C, Burton NW, Khan SR, Uddin R. Association of carbonated soft drink and fast food intake with stress-related sleep disturbance among adolescents: A global perspective from 64 countries. EClinicalMedicine. 2021;31: 100681. doi:10.1016/j.eclinm.2020.100681

10. Purtell KM, Gershoff ET. Fast Food Consumption and Academic Growth in Late Childhood. Clin Pediatr (Phila). 2015;54: 871–877. doi:10.1177/0009922814561742

11. Xu H, Wu X, Wan Y, Zhang S, Yang R, Wang W, et al. Interaction effects of co-consumption of fast food and sugar-sweetened beverages on psychological symptoms: Evidence from a nationwide survey among Chinese adolescents. Journal of Affective Disorders. 2020;276: 104–111. doi:10.1016/j.jad.2020.07.030

12. Asghari G, Yuzbashian E, Mirmiran P, Mahmoodi B, Azizi F. Fast Food Intake Increases the Incidence of Metabolic Syndrome in Children and Adolescents: Tehran Lipid and Glucose Study. PLoS One. 2015;10: e0139641. doi:10.1371/journal.pone.0139641

13. Pereira MA, Kartashov AI, Ebbeling CB, Van Horn L, Slattery ML, Jacobs DR, et al. Fast-food habits, weight gain, and insulin resistance (the CARDIA study): 15-year prospective analysis. Lancet. 2005;365: 36–42. doi:10.1016/S0140-6736(04)17663-0

14. Kucharczuk AJ, Oliver TL, Dowdell EB. Social media’s influence on adolescents’ food choices: A mixed studies systematic literature review. Appetite. 2022;168: 105765. doi:10.1016/j.appet.2021.105765

15. Ashdown-Franks G, Vancampfort D, Firth J, Smith L, Sabiston CM, Stubbs B, et al. Association of leisure-time sedentary behavior with fast food and carbonated soft drink consumption among 133,555 adolescents aged 12-15 years in 44 low- and middle-income countries. Int J Behav Nutr Phys Act. 2019;16: 35. doi:10.1186/s12966-019-0796-3

16. Tambalis KD, Panagiotakos DB, Psarra G, Sidossis LS. Association between fast-food consumption and lifestyle characteristics in Greek children and adolescents; results from the EYZHN (National Action for Children’s Health) programme. Public Health Nutr. 2018;21: 3386–3394. doi:10.1017/S1368980018002707

17. Ma X, Chen Q, Pu Y, Guo M, Jiang Z, Huang W, et al. Skipping breakfast is associated with overweight and obesity: A systematic review and meta-analysis. Obes Res Clin Pract. 2020;14: 1–8. doi:10.1016/j.orcp.2019.12.002

18. Full article: Dietary patterns and associated lifestyle factors among university students in Qatar. [cited 20 Apr 2026]. Available: https://www.tandfonline.com/doi/full/10.1080/07448481.2021.1996374

19. Bencsik P, Lusher L, Taylor RLC. Slow traffic, fast food: The effects of time lost on food store choice. Journal of Urban Economics. 2025;146: 103737. doi:10.1016/j.jue.2025.103737

20. Kabir A, Miah S, Islam A. Factors influencing eating behavior and dietary intake among resident students in a public university in Bangladesh: A qualitative study. PLoS One. 2018;13: e0198801. doi:10.1371/journal.pone.0198801

21. Saeed N, Saleem K, Ashraf R, Aslam M. Factors influencing fast food consumption among university students. Nutrire. 2025;50: 24. doi:10.1186/s41110-025-00330-w

22. Castro O, Bennie J, Vergeer I, Bosselut G, Biddle SJH. How Sedentary Are University Students? A Systematic Review and Meta-Analysis. Prev Sci. 2020;21: 332–343. doi:10.1007/s11121-020-01093-8

23. Roberts CJ, Ryan DJ, Campbell J, Hardwicke J. Self-reported physical activity and sedentary behaviour amongst UK university students: a cross-sectional case study. Critical Public Health. 2024;34: 1–17. doi:10.1080/09581596.2024.2338182

24. Bangladesh University Grants Commission. 50th Annual Report 2023. In: Bangladesh University Grants Commission [Internet]. [cited 20 Apr 2026]. Available: http://ugc.gov.bd/pages/annual-reports/%E0%A7%AB%E0%A7%A6%E0%A6%A4%E0%A6%AE-%E0%A6%AC%E0%A6%BE%E0%A6%B0%E0%A7%8D%E0%A6%B7%E0%A6%BF%E0%A6%95-%E0%A6%AA%E0%A7%8D%E0%A6%B0%E0%A6%A4%E0%A6%BF%E0%A6%AC%E0%A7%87%E0%A6%A6%E0%A6%A8-%E0%A7%A8%E0%A7%A6%E0%A7%A8%E0%A7%A9-93bc29-6922de8f933eb65569e1b95b

25. Deliens T, Clarys P, De Bourdeaudhuij I, Deforche B. Determinants of eating behaviour in university students: a qualitative study using focus group discussions. BMC Public Health. 2014;14: 53. doi:10.1186/1471-2458-14-53

26. Didarloo A, Khalili S, Aghapour AA, Moghaddam-Tabrizi F, Mousavi SM. Determining intention, fast food consumption and their related factors among university students by using a behavior change theory. BMC Public Health. 2022;22: 314. doi:10.1186/s12889-022-12696-x

27. Sajjad M, Bhatti A, Hill B, Al-Omari B. Using the theory of planned behavior to predict factors influencing fast-food consumption among college students. BMC Public Health. 2023;23: 987. doi:10.1186/s12889-023-15923-1

28. Banik R, Naher S, Pervez S, Hossain MdM. Fast food consumption and obesity among urban college going adolescents in Bangladesh: A cross-sectional study. Obesity Medicine. 2020;17: 100161. doi:10.1016/j.obmed.2019.100161

29. Islam IB, Ali MdS, Islam MdR, Islam S, Tabassum N, Rana MdM, et al. Junk Food Consumption Trends, Patterns, and Awareness of Its Health Risks Among University Students in Bangladesh: A Cross-Sectional Study. Health Sci Rep. 2026;9: e71991. doi:10.1002/hsr2.71991

30. Moreira P, Santos S, Padrão P, Cordeiro T, Bessa M, Valente H, et al. Food Patterns According to Sociodemographics, Physical Activity, Sleeping and Obesity in Portuguese Children. Int J Environ Res Public Health. 2010;7: 1121–1138. doi:10.3390/ijerph7031121

31. Watson NF, Badr MS, Belenky G, Bliwise DL, Buxton OM, Buysse D, et al. Recommended Amount of Sleep for a Healthy Adult: A Joint Consensus Statement of the American Academy of Sleep Medicine and Sleep Research Society. Sleep. 2015;38: 843–844. doi:10.5665/sleep.4716

32. Domoff SE, Meers MR, Koball AM, Musher-Eizenman DR. The validity of the Dutch Eating Behavior Questionnaire: some critical remarks. Eat Weight Disord. 2014;19: 137–144. doi:10.1007/s40519-013-0087-y

33. Fletcher EA, McNaughton SA, Crawford D, Cleland V, Della Gatta J, Hatt J, et al. Associations between sedentary behaviours and dietary intakes among adolescents. Public Health Nutr. 2018;21: 1115–1122. doi:10.1017/S136898001700372X

34. Bui C, Lin L-Y, Wu C-Y, Chiu Y-W, Chiou H-Y. Association between Emotional Eating and Frequency of Unhealthy Food Consumption among Taiwanese Adolescents. Nutrients. 2021;13: 2739. doi:10.3390/nu13082739

35. Tareq AM, Mahmud MdH, Billah MdM, Hasan MdN, Jahan S, Hossain MdM, et al. Fast-food and obesity: Status among the young adult population in Bangladesh. Narra J. 2022;2: e86. doi:10.52225/narraj.v2i3.86

